# Impact of COVID-19 Upon Changes in Emergency Room Visits with Chest Pain of Possible Cardiac Origin

**DOI:** 10.1101/2020.08.04.20167908

**Authors:** Adeel A. Butt, Anand Kartha, Nidal Asaad, Aftab M. Azad, Roberto Bertollini, Abdul-Badi Abou-Samra

**Author notes:** Address all correspondence to: Adeel A. Butt, MBBS, MS, FACP, FIDSA, PO Box 3050, Department of Medicine, Hamad Medical Corporation, Doha, Qatar.

## Abstract

**Background:** A decrease in Emergency Department (ED) visits for cardiac conditions has recently been reported from the US and Western Europe due to the COVID-19 pandemic. The data are still scant, and the correlation between cardiac symptoms and confirmed diagnoses are not available. There are no reports on changes in ED volumes at a national level, or from countries in the Asia-Middle Eastern region.

**Methods:** We report data from national referral centers for tertiary care and cardiac care centers in Qatar, which see >80% of cardiac emergencies in the country.

**Results:** We analyzed 102,033 ED visits in the COVID-era (March-April 2020 and 2019) and determined the proportion presenting for cardiac symptoms and their confirmed diagnoses. We observed a 16-37% decline in ED volumes overall, with a 25-50% decline in patients presenting with cardiac symptoms in March-April 2020 compared with March-April 2019. Among those presenting with cardiac symptoms, we observed a 24-43% decline in cardiac diagnoses.

**Conclusions:** A sharp decline in patients presenting with cardiac symptoms was observed in the COVID-era. A post-COVID surge in patients with these conditions may be anticipated and preparations should be made to address it.

## Background

Earlier anecdotal reports of a decline in acute care hospitalizations during the current COVID-19 pandemic have been confirmed with recent publications reporting a decline in overall admissions and admission for acute coronary syndrome.^1–6^ These reports have all been from the US or Western Europe with relative homogenous populations. Data from countries with large migrant worker and more diverse populations are not available. Changes in volumes and visits to the Emergency Department (ED) for cardiac symptoms, and their correlation with final diagnosis in the COVID-era compared with pre-COVID era are also largely unknown. Our aim was to determine the change in volumes of patients presenting to large Emergency Departments with cardiac symptoms before and after the current COVID-19 pandemic, and to determine the proportion of a confirmed cardiac diagnosis among them.

## Methods

In the State of Qatar, over 80% of patients with suspected acute coronary events are seen in two hospitals: Hamad General Hospital (HGH), which is the nation’s largest tertiary care referral center and the Heart Hospital (HH), which is the only specialty hospital designated specifically for cardiac care. Both hospitals are accredited by the Joint Commission International and utilize the same electronic health records system (Cerner®). Patients retain the same unique medical record number across both hospitals. The first COVID-19 patient in Qatar was diagnosed on February 29, 2020. We determined the number of individuals presenting to the EDs at the above mentioned hospitals during March-April 2020 (COVID-era) and compared it with the numbers in March-April 2019 (pre-COVID-era). At presentation, a trained nurse obtained the history of presenting complaint. Patients with acute chest pain with characters suggestive of cardiac origin (e.g. radiation to the left arm, other concomitant symptoms), shortness of breath (except when with cough and fever), palpitations and syncope/near syncope were categorized as having cardiac symptoms by three physicians among the authors (AAB, AK, AA), who independently reviewed all presenting complaints and assigned this category.

## Results

A total of 102,033 ED visits were recorded over the study period. Compared with the same month in 2019, a 16.2% decline in total ED visits was noted in March 2020 and a 37.4% decline in April 2020. **(Table)** A similar decline in patients presenting with cardiac symptoms was also noted, with a 25.0% decline in March 2020 and a 49.9% decline in April 2020 compared with the same month in 2019. Among those presenting with cardiac symptoms, a sharp decline in confirmed cardiac diagnoses was also noted (24.3% in March 2020; 42.6% in April 2020). A list of all patients with their presenting symptoms, the breakdown of cardiac symptoms and confirmed diagnoses among those with cardiac symptoms are provided in **supplementary tables 1, 2 and 3** respectively.

**Table.** Total Emergency Department visits for cardiac symptoms (chest pain likely cardiac, palpitations, shortness of breath and syncope/near syncope) and a breakdown of confirmed cardiac diagnoses among those with cardiac symptoms in months after and before COVID-19 cases.

|  | 2019-03 |  | 2019-04 |  | 2020-03 |  | 2020-04 |  |
| --- | --- | --- | --- | --- | --- | --- | --- | --- |
| **Total ED visits** | **29,908** |  | **28,950** |  | **25,049** |  | **18,126** |  |
| % change from previous year | -- |  | -- |  | -16.2% |  | -37.4% |  |
|  | N | % | N | % | N | % | N | % |
| **Total ED visits for cardiac symptoms** | **3,185** | **31.9%** | **2,941** | **29.4%** | **2,389** | **23.9%** | **1,472** | **14.7%** |
| % change from previous year | -- | -- | -- | -- | -25.0% |  | -49.9% |  |
| **Cardiac diagnoses among those with cardiac symptoms** | **445** | **33.4%** | **350** | **26.3%** | **337** | **25.3%** | **201** | **15.1%** |
| % change from previous year | -- | -- | -- | -- | -24.3% |  | -42.6% |  |
| Acute coronary syndrome | 191 | 6.0% | 167 | 5.7% | 170 | 7.1% | 113 | 7.7% |
| Angina or coronary artery disease without acute coronary syndrome | 93 | 2.9% | 74 | 2.5% | 32 | 1.3% | 7 | 0.5% |
| Arrhythmia | 78 | 2.4% | 42 | 1.4% | 47 | 2.0% | 26 | 1.8% |
| Cardiac arrest | 20 | 0.6% | 14 | 0.5% | 20 | 0.8% | 19 | 1.3% |
| Congestive heart failure | 62 | 1.9% | 47 | 1.6% | 63 | 2.6% | 35 | 2.4% |
| Valvular Disease | 1 | 0.0% | 6 | 0.2% | 5 | 0.2% | 1 | 0.1% |

1 Denominator for percentage calculation is the total number of ED visits for that month
2 Denominator for percentage calculation is the total number of ED visits for cardiac symptoms

## Discussion

To our knowledge, this is the first study to provide detailed information on cardiac symptoms and confirmed cardiac diagnoses in the COVID-era compared with pre-COVD-era. The sharp decline in overall ED visits confirms anecdotal reports and recently published reports from regional hospitals and healthcare systems in the US and Western Europe. The actual number and proportion of patients presenting with cardiac symptoms also dropped sharply, as did the number and proportion of persons with a confirmed cardiac diagnosis among those presenting with cardiac symptoms. These trends are worrisome since acute cardiac conditions generally require immediate intervention and appropriate emergency and/or inpatient care for achieving optimal outcomes. The reasons for these declines are most likely related to the travel and movement restrictions in the COVID-era, since all other parameters for patient access, flow and care remained unchanged. These declines may be among patients with serious conditions who were otherwise unable to travel to an ED, or a self-conclusion among patients considering the symptoms to be non-emergent in nature. In either scenario, a surge of patients may be expected with relaxation of restriction of movement and appropriate steps need to be taken in anticipation of such a surge. It is also critical to determine any increase in cardiac related mortality in the community and institute interventions to mitigate such risk.

## Conclusions

In conclusion, a sharp decline in total ED visits, as well as visits for cardiac symptoms was observed at a national level, with a parallel decrease in number and proportion of persons with a confirmed cardiac diagnosis. A post-COVID surge in patients with these conditions may be anticipated and preparations should be made to address it.

## Data Availability

Data are not publicly available.

## Abbreviations

ED: Emergency Department
HGH: Hamad General Hospital
HH: Heart Hospital

## Declarations

### Ethics approval and consent to participate

The study was approved by the Institutional Review Board of Hamad Medical Corporation. Waiver of informed consent was granted by the Hamad Medical Corporation Institutional Review Board.

### Consent for publication

All authors provided consent for publication. There are no patient or subject data that require consent.

### Availability of data and material

Aggregate data may be made available through a forma request to Hamad Medical Corporation, who will make any and all decisions regarding data sharing. No personally identifiable data will be provided.

### Authors’ contributions

Concept, design, data acquisition, funding: AAB Data analysis: AAB, AK, AA

Data interpretation: AAB, AK, NA, AMA, RB, AA

Critical appraisal and important intellectual content: AAB, AK, NA, AMA, RB, AA All authors have read and approve the manuscript.

## Acknowledgements

None

## Additional Files

File Name: Supplementary Data

Title of data: Supplementary tables

Description of data:

Supplementary table 1. Chief complaints of patients presenting to the Emergency Departments at all participating hospitals over the study period (listed alphabetically).

Supplementary table 2. Cardiac symptoms.

Supplementary table 3. Confirmed diagnoses for those presenting with cardiac symptoms

## References

1. Bernstein LS, F.S.;. Patients with heart attacks, strokes and even appendicitis vanish from hospitals. https://www.washingtonpost.com/health/patients-with-heart-attacks-strokes-and-even-appendicitis-vanish-from-hospitals/2020/04/19/9ca3ef24-7eb4-11ea-9040-68981f488eed_story.html. Washington Post April 19, 2020.

2. De Filippo O, D’Ascenzo F, Angelini F, et al. Reduced Rate of Hospital Admissions for ACS during Covid-19 Outbreak in Northern Italy. N Engl J Med 2020. 10.1056/NEJMc2009166.

3. Baldi E, Sechi GM, Mare C, et al. Out-of-Hospital Cardiac Arrest during the Covid-19 Outbreak in Italy. N Engl J Med 2020. 10.1056/NEJMc2010418.

4. Solomon MD, McNulty EJ, Rana JS, et al. The Covid-19 Pandemic and the Incidence of Acute Myocardial Infarction. New England Journal of Medicine 2020. DOI 10.1056/NEJMc2015630.

5. Baum A, Schwartz MD. Admissions to Veterans Affairs Hospitals for Emergency Conditions During the COVID-19 Pandemic. JAMA 2020. DOI 10.1001/jama.2020.9972.

6. Garcia S, Albaghdadi MS, Meraj PM, et al. Reduction in ST-Segment Elevation Cardiac Catheterization Laboratory Activations in the United States during COVID-19 Pandemic. J Am Coll Cardiol 2020. 10.1016/j.jacc.2020.04.011.

7. Wolfler A, Mannarino S, Giacomet V, Camporesi A, Zuccotti G. Acute myocardial injury: a novel clinical pattern in children with COVID-19. The Lancet Child & Adolescent Health.

